# Intraoperative OCT-Guided Pneumodescemetopexy and Corneal Compression Sutures for Extensive Acute Corneal Hydrops

**DOI:** 10.64898/2026.04.15.26350813

**Authors:** Ioannis Giachos, Ahmed Hamdy Oreaba, Usama Kanj, Saba Anwar, Rupinder Chahal, Anil Aralikatti, Darren S. J. Ting

## Abstract

**Purpose:** To highlight the roles of intraoperative optical coherence tomography (iOCT) in managing acute corneal hydrops (ACH) and outcomes of iOCT-guided pneumodescemetopexy and corneal compression sutures.

**Methods:** This was a retrospective, consecutive, interventional case series of patients with keratoconus who presented with significant ACH and underwent iOCT-guided pneumodescemetopexy (18% sulfur hexafluoride gas) and compression sutures at Birmingham and Midland Eye Centre, UK, between Aug 2023 and May 2025.

**Results:** Five patients were included; mean age was 32.3±6.6 years old and 3 (60%) were male. The mean follow-up duration was 16.3±5.6 months. At presentation, the mean corrected-distance-visual-acuity (CDVA) was 1.90±0.67 logMAR, central corneal thickness (CCT) was 1187.6±372.6μm, maximal corneal thickness was 1624.0±383.5μm and maximal height and diameter of pre-Descemet layer/Descemet membrane (PDL/DM) detachment was 1014.6±366.4μm and 4456.0±839.4μm, respectively. The surgery successfully achieved complete PDL/DM attachment in all cases, with a mean time from surgery to ACH resolution of 17.8±8.0 days. iOCT successfully visualized the area of PDL/DM break/detachment, revealed the involvement of PDL (evidenced by a persistent taut type 1 DM detachment after gas tamponade), and guided the placement of compression sutures. Compared to preoperative, there was a significant improvement in the mean CDVA (0.52±0.32 logMAR; p=0.014) at last follow-up. One patient required a repeat procedure to fully attach the PDL/DM.

**Conclusions:** This study demonstrated favorable outcomes of iOCT-guided pneumodescemetopexy and corneal compression sutures. iOCT revealed the involvement of PDL in ACH and provided plausible explanations why pneumodescemetopexy alone may not be able to resolve significant ACH rapidly in certain cases.

## INTRODUCTION

Keratoconus is a bilateral progressive corneal ectatic disorder characterized by cone-like steepening of the cornea. It typically presents during puberty or early adulthood, and if left untreated, it can gradually progress and lead to significant vision loss.^1,2^ It is a multifactorial disease, with eye rubbing, allergic eye disease, and genetic factors playing important roles.^3,4^

The presentation and rate of progression of keratoconus can be highly variable. The innovation of corneal crosslinking has helped stabilize the progression of keratoconus, preserved the vision, and reduced the need for keratoplasty in the past 1-2 decades.^5^ However, keratoplasty may still be warranted in patients with moderate-to-severe cases or those who cannot tolerate contact lens.^6–8^ Rarely, some patients may develop acute corneal hydrops (ACH) as a complication of severe keratoconus.

ACH is an uncommon yet sight-threatening complication of corneal ectatic diseases, affecting around 0.2-2.8% of patients with keratoconus.^9^ It can also present in other conditions like post-LASIK ectasia, penetrating keratoplasty, and post-CXL.^10–13^ ACH is often managed conservatively with topical treatment, including steroids, hypertonic sodium chloride, antibiotic, cycloplegics, intraocular pressure-lowering drops, and/or bandage contact lens, though it may take 2-6 months or even longer with conservative management.^10^ In severe cases, ACH is usually unresponsive to medical treatment and necessitate surgical treatment.

The choice of surgical intervention is largely dependent on the disease severity of ACH and the primary objective of the surgery.^10^ Surgical interventions can range from pneumodescemetopexy / intracameral air/gas tamponade and corneal compression sutures (to approximate the Descemet membrane [DM] break and detachment) to various forms of keratoplasty, including endothelial keratoplasty (to replace the affected DM), modified deep anterior lamellar keratoplasty (DALK), predescemetic DALK, and penetrating keratoplasty (to address edematous cornea and scarring).^10,14,15^ While pneumodescemetopexy serves as a less invasive procedure and could expedite the resolution of ACH, studies have shown that ACH may still persist for 2-3 months after intracameral gas injections and may require subsequent keratoplasty to resolve the condition or improve the vision.^10,16,17^

Intraoperative optical coherence tomography (iOCT) is a useful technology for real-time intraoperative imaging of ocular tissues and guiding various ocular surgeries, including corneal transplantation, cataract and refractive surgeries, and vitreoretinal surgery.^18,19^ In this study, we aimed to examine the effectiveness and safety of iOCT-guided pneumodescemetopexy and corneal compression sutures for extensive ACH. In addition, we aimed to demonstrate the roles of iOCT in guiding the surgical management and revealing the involvement of pre-Descemet’s layer/Dua’s layer (PDL) in the pathogenesis of ACH. This study also provided plausible explanations as to why pneumodescemetopexy (using either intracameral air or gas tamponade) alone may not be able to resolve extensive ACH rapidly in certain cases.

## METHODS

This was a retrospective, consecutive, interventional case series of five patients with keratoconus who presented with extensive ACH and underwent surgical treatment at Birmingham and Midland Eye Centre (BMEC), UK, between Aug 2023 and May 2025. This study was approved as a clinical audit by the local Clinical Effectiveness Team at Sandwell and West Birmingham NHS Trust, Birmingham, UK (Reference: #3118). Informed consent was obtained from all included patients for the treatment and publication. All data have been de-identified prior to use in this study.

All patients underwent slit-lamp biomicroscopic examination/photography and anterior segment optical coherence tomography (AS-OCT; Heidelberg Anterion system, Germany), which demonstrated significant corneal edema with DM detachment and break in all cases, confirming the diagnosis of ACH.^19^ Relevant data, including preoperative and postoperative corrected-distance-visual-acuity (CDVA), central corneal thickness (CCT), maximal corneal thickness (MCT), maximal height and diameter of the DM detachment, the location of the tear, and time to complete resolution of edema, were recorded and analyzed. CCT was defined as the distance between the corneal epithelium and the DM (detached or attached) at the central cornea, whereas MCT (i.e., maximal perpendicular distance between the epithelium and the detached DM), maximal height of DM detachment (i.e., maximal perpendicular distance between the posterior corneal stroma and the DM) and maximal diameter of DM detachment (i.e., maximal horizontal distance between the two ends of the posterior stroma demarcated by the DM detachment) were measured at the area with maximal DM detachment (**Figure 1**).

**Figure 1.**
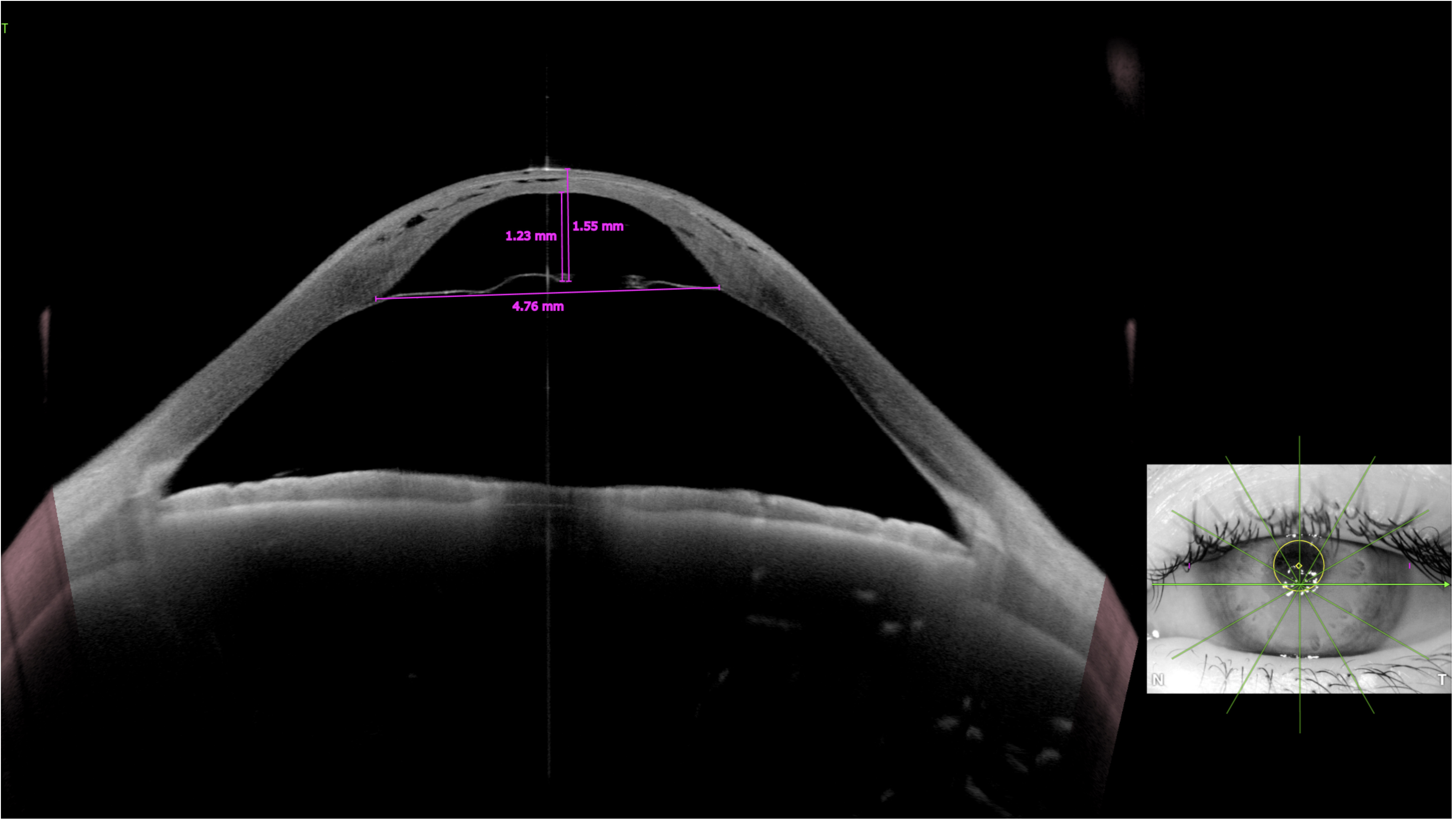
AS-OCT showing a taut, hyper-reflective line demarcating the break at the pre-Descemet’s layer / Descemet membrane (PDL/DM), with resultant significant corneal edema. Maximal corneal thickness, height and diameter of DM detachment was assessed manually at the maximal PDL/DM detachment site. The maximal height of DM detachment is defined as the maximal perpendicular distance between the posterior corneal stroma and the detached DM, whereas the maximal diameter of DM detachment is defined by the maximal horizontal distance between the two ends of the posterior stroma demarcated by the DM detachment. Central corneal thickness was also measured manually (not shown in this picture).

The main outcome measures of this study were the success rate of complete anatomical reattachment of DM/PDL and the time from surgery to resolution of ACH, whereas the secondary outcome measure was the CDVA.

### Surgical technique

Informed consent was obtained from all patients prior to the procedure. All patients underwent pneumodescemetopexy [with iso-expansile concentration (18%) of sulfur hexafluoride (SF_6_) gas] and corneal compression sutures guided by iOCT within four weeks of initial presentation of ACH (except for patient 4). All cases were performed by a corneal consultant, either DSJT (n=4) or AA (n=1), in the operating room under sub-Tenon local anesthesia or general anesthesia (on patient request).

Briefly, a paracentesis was created to release a small amount of aqueous humor. A 30-gauge cannula mounted on a 2Lml syringe was used to inject 18% SF_6_ gas through the same paracentesis into the anterior chamber to obtain a full gas fill. Despite a fully filled anterior chamber, the PDL/DM break and detachment failed to attach completely to the posterior stroma. This was evidenced by a taut hyper-reflective line on the iOCT, which disappeared on corneal compression by a blunt cannula over the PDL/DM break and re-appeared upon the discontinuation of compression (**Figure 2**). Further injection of gas bubble was proven to be ineffective to attach the PDL/DM, as demonstrated by iOCT, as the gas escaped through the PDL/DM break into the area between the posterior stroma and the detached PDL/DM, forming a “double chamber”. As a result, 3-4 full-thickness corneal compression sutures were placed perpendicular to the PDL/DM break to completely attach the PDL/DM (confirmed by iOCT). At the end of the procedure, the anterior chamber was left with 60-70% gas fill, and subconjunctival injections of cefuroxime and betamethasone and oral acetazolamide 500mg were given. Patients were kept supine posture for 3-4 hours postoperatively and then as much as possible for the subsequent 2 days. Snapshots of various stages of the operation were illustrated in **Figure 2**. The surgical technique and the key iOCT findings described in this study is highlighted in **Video 1**.

**Figure 2.**
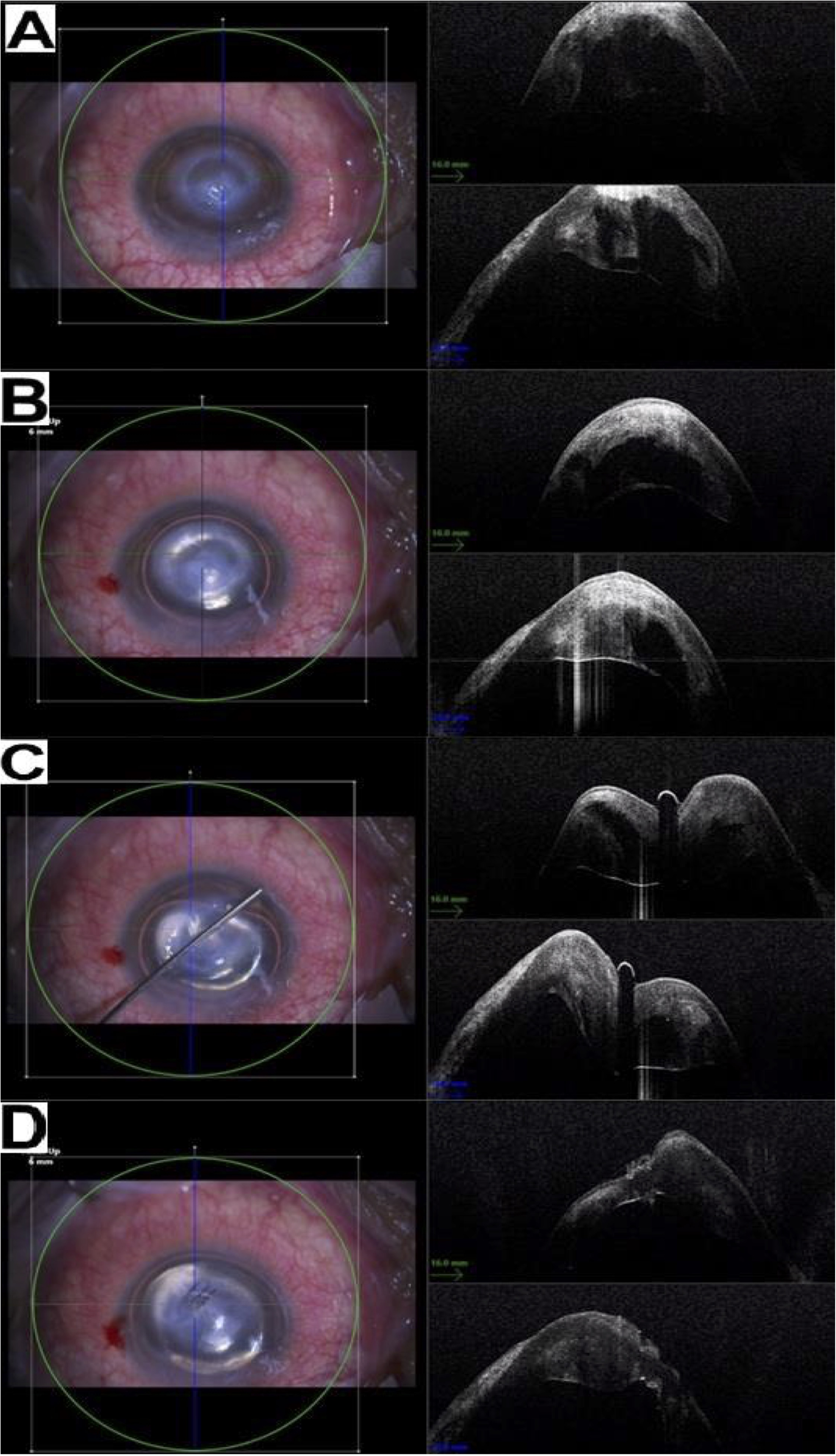
Snapshots of surgical video with intraoperative optical coherence tomography (iOCT) demonstrating the intraoperative steps of pneumodescemetopexy and corneal compression sutures for managing extensive acute corneal hydrops (ACH). **(A)** iOCT demonstrating extensive central ACH with corneal edema and intrastromal cleft. **(B)** After intracameral injection of 18% SF_6_ gas into the anterior chamber, the pre-Descemet’s layer/Descemet membrane (PDL/DM) detachment persisted, evidenced by the taut, hyper-reflective line at the posterior cornea (type 1 DM detachment). There was also some gas escaped into the gap between the posterior stroma and the detached PDL/DM, inhibiting PDL/DM attachment. **(C)** External compression of the cornea with a Rycroft cannula pushed the air bubble out from the space between posterior stroma and detached PDL/DM, and enabled the attachment of PDL/DM to the posterior stroma. **(D)** Attachment of PDL/DM to the posterior stroma was achieved with three corneal compression sutures and pneumodescemetopexy.

All patients were examined at 1 day, 1 week, and 4 weeks postoperative then 1-2 monthly thereafter. Sutures were removed at 4-6 weeks postoperative, unless patients requested to leave them in situ. All patients received topical preservative-free levofloxacin 0.5% 4 times a day for 1 month, dexamethasone 0.1% 4 times a day for 1 month, hypertonic saline 5% 4 times a day for 1 month, and cyclopentolate 1% 3 times a day for 1 week. Slit-lamp biomicroscopic examination (and/or photography) and ASOCT were performed at each visit to monitor and confirm the resolution of ACH. All included patients completed at least 6 months postoperative follow-up.

## RESULTS

A total of five patients with keratoconus were included in this study; the mean age was 32.3±6.6 years old and 3 (60%) were male. The mean follow-up duration was 16.4±5.6 months. Demographic factors, clinical details, preoperative and postoperative vision, and time taken for complete resolution of corneal edema from initial presentation are detailed in **Table 1**. None of the patients had a history of CXL.

At presentation, the mean corrected-distance-visual-acuity (CDVA) was 1.90±0.67 logMAR, CCT was 1187.6±372.6μm, MCT was 1624.0±383.5μm and maximal height and diameter of DM detachment was 1014.6±366.4μm and 4456.0±839.4μm, respectively. The mean time from the onset of ACH to the initial presentation was 2.2±1.6 days, and the mean time from the initial presentation to surgical intervention was 18.2±18.4 days. All patients had the surgery within 4 weeks of the onset of hydrops, except for patient #4, who had the surgery at 52 days after the onset of hydrops (due to the delay in referral from the initial eye emergency team to the corneal team). Preoperative AS-OCT characteristics of ACH and the tear are provided in **Figure 3**. All tears were shown to involve either the central 3 mm area of the cornea (3, 60%) or the inferior paracentral cornea (2, 40%), with a type 1 DM detachment (evidenced by the presence of a hyper-reflective taut line on AS-OCT).

**Figure 3.**
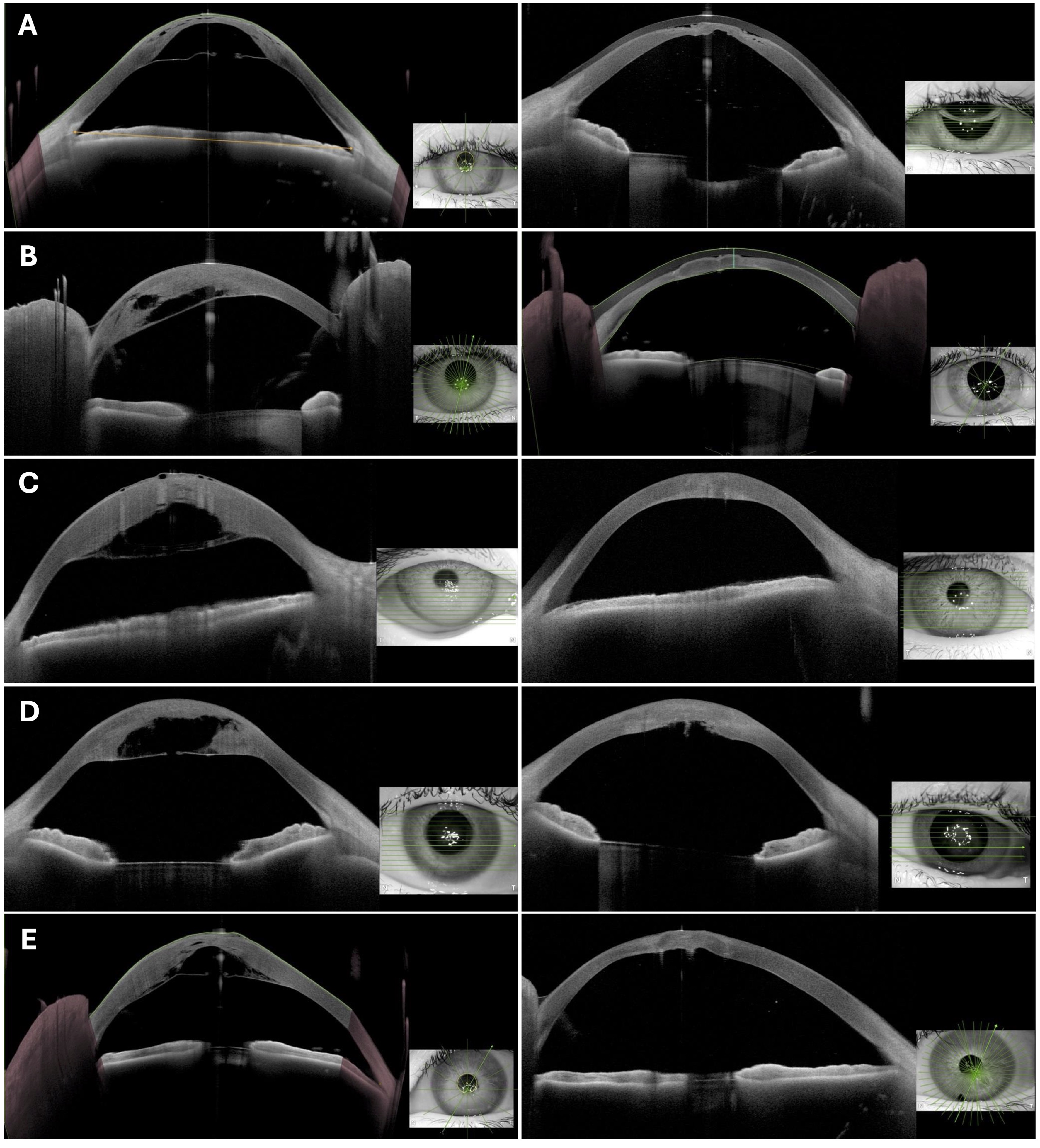
Preoperative (left column) and postoperative (right column) anterior segment optical coherence tomography (AS-OCT) images of all patients. (A) Patient #1: Preop vs. postop (day 7); (B) Patient #2: Preop vs. postop (day 11); (C) Patient #3: Preop vs. postop (day 29); (D) Patient #4: Preop vs. postop (day 8 post-repeated surgery); and (E) Patient #5: Preop vs. postop (day 7).

The surgery successfully achieved a complete reattachment of PDL/DM in all cases, with a mean time from surgery to resolution of corneal edema of 17.8±8.0 days. All operated eyes had an improvement in CDVA postoperatively, with a mean improvement of 1.38±0.65 logMAR. Three (60%) patients achieved a CDVA of ≤0.30 logMAR (i.e. Snellen vision of 20/40 or better). The gas bubble completely disappeared from the anterior chamber after 7 days post-injection. Compared to preoperative, significant improvement in the mean CDVA (0.52±0.32 logMAR; p=0.014) and CCT (475.6±123.3μm; p=0.029) was observed at last follow-up.

Intraoperatively, the iOCT successfully facilitated the precise visualization of the area of DM break and detachment. It also revealed a type 1 DM detachment and highlighted the involvement of PDL in ACH in all cases, evidenced by a taut hyper-reflective line at the posterior cornea, even in the presence of a full gas-filled anterior chamber. The findings provided plausible explanation as to why pneumodescemetopexy alone might not be able to resolve extensive ACH rapidly in certain cases.

One patient required a repeat procedure to fully attach the PDL/DM, and one patient required an additional top-up of intracameral air injection at day 1 postoperative to address a minor corneal wound leak at the suture site, which resolved without any further issues. All eyes were comfortable after the resolution of ACH and obviated the need for subsequent optical keratoplasty. Representative preoperative and postoperative slit-lamp photographs and AS-OCT images are provided in **Figure 3 and Figure 4**.

**Figure 4.**
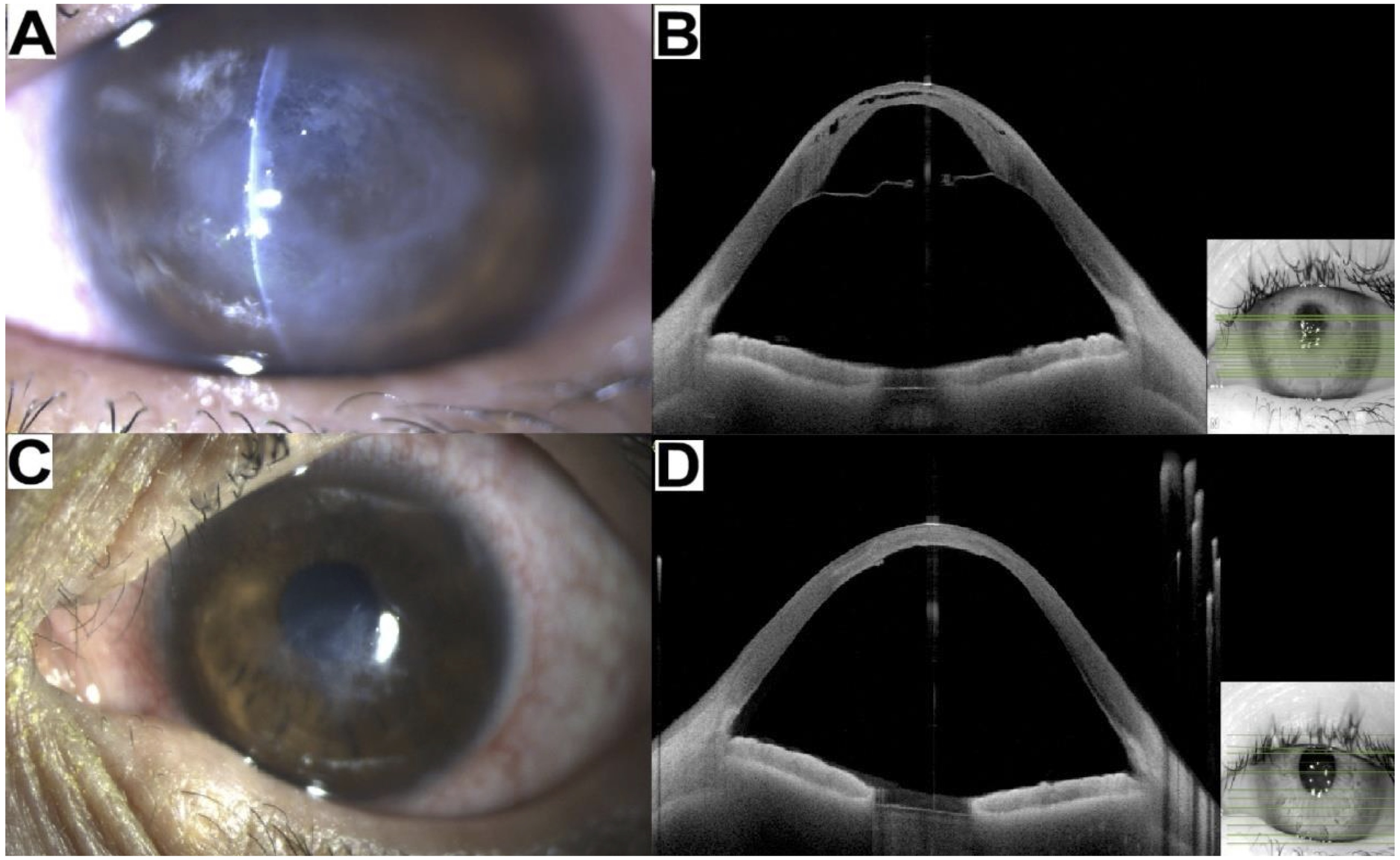
A case of extensive acute corneal hydrops (ACH) in a patient with keratoconus (Patient #1). **(A-B)** Slit-lamp photography and anterior segment optical coherence tomography (AS-OCT) demonstrating an extensive central ACH with a type 1 pre-Descemet’s layer/Descemet membrane (PDL/DM) detachment, significant corneal edema and intrastromal cleft. **(C-D)** Postoperative slit-lamp photography and AS-OCT demonstrating complete resolution of corneal edema and attachment of PDL/DM with mild corneal scarring at 6 weeks after intraoperative OCT-guided pneumodescemetopexy and corneal compression sutures (which were already removed).

## DISCUSSION

In this study, we demonstrated that extensive ACH can be effectively managed with iOCT-guided pneumodescemetopexy and corneal compression sutures, with all treated eyes achieving a complete anatomical reattachment of PDL/DM, rapid resolution of corneal edema with limited corneal scarring, and improvement in CDVA. More importantly, our study highlighted the roles of iOCT in guiding the surgical management of ACH and supporting the involvement of PDL in the pathogenesis of ACH.

iOCT has emerged as a valuable tool for guiding ophthalmic surgeons during various types of eye surgeries, including anterior and posterior segment surgeries. Within the field of keratoplasty, iOCT has demonstrated its value in deep anterior lamellar keratoplasty (DALK) and Descemet membrane endothelial keratoplasty (DMEK). During DALK, iOCT can visualize the depth of trephination and the depth of cannula placement as well as the formation of big bubble.^20,21^ Similarly, decision making in DMEK has been greatly augmented as the graft insertion, its orientation and any residual interface fluid can be observed and dealt with accordingly.^22–24^ Moreover, iOCT has been beneficial to posterior segment surgeons as it can guide the peeling of internal limiting membrane (during macular hole surgery) and epiretinal membrane, delamination in proliferative diabetic retinopathy, and sub-retinal injections (e.g. for retinal gene therapy).^25^

In this study, we made three clinically meaningful observations. First, we showed that iOCT-guided pneumodescemetopexy and corneal compression sutures were able to achieve rapid resolution of extensive ACH (mean height of PDL/DM detachment of 1014.6±366.4μm), with a mean resolution time of 17.8±8.0 days. iOCT enables visualization of the area and extent of PDL/DM detachment, informing the precise placement and the required number of the full-thickness corneal compression sutures. It also helps confirming the attachment of PDL/DM after the insertion of compression sutures. While visualization of the DM/PDL break is possible with the use of intracameral air/gas bubble, gleaned from our personal experience and findings of Ashena et al.,^26^ the use of iOCT provides a real-time view of the affected area as well as the end result following the surgical intervention. Other studies have also reported the use of iOCT for guiding the drainage of intrastromal fluid pockets and pneumodescemetopexy (with and without compression sutures) in eyes with ACH.^27,28^

Previous studies have shown that ACH typically resolves within 2-6 months of the onset with conservative management, but may persist even more than 6 months particularly in those with severe ACH.^10,16,29^ This can result in prolonged ocular discomfort, corneal scarring, vascularization, and reduced vision.^29–31^ Moreover chronically detached PDL/DM is populated with keratocyte-derived cells, leading to fibrosis and eventual failure to reattach.^32^ The rapid resolution of corneal edema observed in our study parallels the findings of several other studies, which reported an average resolution time of 1-3 weeks after pneumodescemetopexy [using 14% perfluoropropane (C3F8)] and corneal compression sutures.^33,34^ The rapid resolution of ACH also helped reduce the risk of corneal scarring, evidenced by the improvement in CDVA in all our included patients. Interestingly, a study has shown that corneal compression sutures alone could also achieve rapid resolution of ACH.^26^ One of our patients (patient #4) required a repeat surgery to fully resolve the ACH, and this might be related to the longer time interval between the onset of hydrops and the surgery (52 days) as chronically detached DM/PDL may lead to fibrosis and increased difficulty in re-attachment.

Another potential benefit of this surgical approach is that it is less invasive than keratoplasty, which is known to be associated complications (e.g. graft infection, rejection, and failure) and requires long-term regular follow-up. However, it is important to bear in mind that pneumodescemetopexy with corneal compression sutures may induce focal suture-related stromal scarring and does not address the underlying corneal ectasia and high irregular astigmatism that is often observed in keratoconic eyes that develop ACH. This was evident in one of our patients (patient #3) who had a successful reattachment of PDL/DM following the surgery but only achieved a CDVA of 20/200 (the presenting vision was counting fingers). However, the patient was happy to be managed conservatively after the operation and declined any further keratoplasty Therefore, careful preoperative patient counselling is important to manage patient expectation. On the other hand, novel techniques such as pre-descemetic DALK could offer rapid resolution of corneal oedema, removal of stromal scarring, correction of high astigmatism, improvement in vision, and mitigate the risk of endothelial graft rejection (as opposed to penetrating keratoplasty), though long-term follow-up is required to monitor for postoperative graft-related complications.^15^ Another common approach for treating severe ACH with visually significant corneal scarring is optical penetrating keratoplasty, which can address the irregular/high astigmatism, stromal scarring and endothelial dysfunction, though it is associated with the risk of suture-related complications, infectious keratitis, and endothelial graft rejection.^36^

Second, the use of iOCT helps reveal the involvement of PDL in the pathogenesis of ACH. This is supported by the intraoperative observation of a taut, hyper-reflective line at the posterior cornea in all our cases, even in the presence of a full gas-filled anterior chamber, resembling a type 1 DM detachment (where the detachment occurs between the posterior stroma and PDL/DM).^35^ If the detachment only involves the DM, the full gas tamponade in the anterior chamber would have attached the DM to the posterior cornea. Conventionally, ACH is believed to be caused by a split in the DM and corneal endothelial layer due to excessive corneal protrusion and thinning, with subsequent influx of aqueous humor in the cornea causing corneal edema and/or intrastromal cleft or cyst.^37^ However, recent studies have provided supportive evidence on the involvement of PDL in the pathogenesis of ACH.^38–40^

PDL is an acellular layer that is predominantly composed of collagen type I, collagen type IV and a high density of elastin (which confers its biomechanical and tensile strength), with bursting pressures up to 700mmHg being demonstrated.^32,41^ Studies have shown that PDL/DM detachment (type 1 DM detachment) behaves differently from pure DM detachments (type 2 DM detachment).^35^ PDL/DM detachment appears as a straight taut, hyperreflective line on AS-OCT, while pure DM detachment is seen as a thin, undulating, double contour hyper-reflective line.^35^ Moreover, the amount of scrolling in a PDL/DM detachment is much less than in a pure DM detachment.

Based on the AS-OCT findings, the characterization of PDL and the observation that postoperative DM detachment alone does not cause ACH, it was proposed that ACH was secondary to a tear in both DM and the PDL, in the context of abnormal collagen and proteoglycan matrix in ectatic corneas.^39,41^ The hypothesis was supported by the failure of inducing ACH in non-keratoconic human donor sclero-corneal discs after a posterior deep 3 mm incision through the DM, PDL and posterior stroma.^39,41^ The involvement of PDL in ACH was similarly suggested by Parker et al.,^38^ who observed that ACH occurs only when both PDL and DM are perforated, but not when DM alone is affected.

Third, iOCT also revealed that a full gas tamponade in the anterior chamber failed to oppose the detached PDL/DM to the posterior stroma in all our cases. In fact, the gas could escape into the gap between the posterior stroma and the detached PDL/DM in certain cases. This interesting *in vivo* finding provides plausible explanation as to why pneumodescemetopexy alone may not be able to resolve severe ACH rapidly in certain cases. For instance, Basu et al.^16^ demonstrated that patients with ACH who underwent pneumodescemetopexy with 14% C3F8 had a mean time to resolution of corneal edema of 78.7±53.2 days. This is significantly slower than patients with ACH that had combined pneumodescemetopexy and compression sutures, as shown in our study (mean time of resolution of 17.8±8.0 days) and other studies. Another study similarly showed that some patients may require up to 3 injections into the anterior chamber to manage ACH.^42^ As a result, the use of corneal compression sutures, perpendicular to the DM break, in addition to pneumodescemetopexy has been proposed. The rationale is that the corneal sutures compress the stroma and subsequently closes the gap between posterior stroma and detached DM, which helps resolve the ACH more effectively and efficiently.^33,34,40,43^

One of the study limitations included a small sample size (due to the disease rarity) and a lack of control group (e.g. ACH without surgical intervention or iOCT-guided surgery). However, the rapid resolution of severe ACH observed in all our included cases and a much faster resolution time than non-treated ACH reported in the literature (around 2-6 months or even longer) provided strong supportive evidence of this surgical approach. We acknowledged that the use of iOCT is not indispensable in the management of severe ACH, but it can facilitate precise visualization of the extent of the DM/PDL break and the successful closure of the break. Moreover, it demonstrated the involvement of PDL and its significance in the management of severe ACH.

In conclusion, this study demonstrated that iOCT serves as a valuable tool in guiding the surgical management of extensive ACH and revealing the involvement of PDL in ACH. It also provides explanation as to why pneumodescemetopexy alone may not be able to resolve severe ACH rapidly, necessitating the need for corneal compression sutures in some cases.

## Data Availability

All data produced in the present work are contained in the manuscript.

## VIDEO LEGEND

**Video 1.** Surgical video demonstrating the technique of intraoperative optical coherence tomography (iOCT)-guided pneumodescemetopexy and corneal compression sutures for managing extensive acute corneal hydrops. The video also reveals a type 1 Descemet membrane detachment (DMD) and the involvement of pre-Descemet’s layer (PDL) in the pathogenesis of acute corneal hydrops and explains why pneumodescemetopexy alone may not fully resolve extensive acute corneal hydrops in some cases.

