## Supplementary material for "Intraoperative OCT-Guided Pneumodescemetopexy and Corneal Compression Sutures for Extensive Acute Corneal Hydrops": Table 1

**Table 1.** Summary of patients with extensive acute corneal hydrops (ACH) who underwent pneumodescemetopexy and corneal compression sutures.

| **Patient** | **Age** | **Gender** | **Laterality** | **Presenting CDVA, logMAR** | **Presenting CCT, μm** | **Time from onset to initial presentation, days** | **Time from presentation to surgery, days** | **Location of ACH and tear** |
| --- | --- | --- | --- | --- | --- | --- | --- | --- |
| 1 | 30s | Male | Left eye | 2.30 | 1470 | 5 | 3 | Central 3 mm |
| 2 | 40s | Female | Right eye | 0.78 | 844 | 3 | 26 | Inferior paracentral |
| 3 | 30s | Male | Right eye | 1.70 | 1770 | 1 | 6 | Central 3 mm |
| 4 | 20s | Male | Left eye | 2.80 | 1030 | 1 | 51 | Inferior paracentral |
| 5 | 30s | Female | Left eye | 1.90 | 824 | 1 | 5 | Central 3 mm |

**Table 1 (Cont).** Summary of patients with extensive acute corneal hydrops (ACH) who underwent pneumodescemetopexy and corneal compression sutures.

| **Patient** | **MCT at detachment site, μm** | **Max. height PDL/DM detachment, μm** | **Max. diameter of PDL/DM detachment, μm** | **Number of sutures** | **Final CDVA, logMAR** | **Final CCT. μm** | **Time to resolution of corneal edema, days** |
| --- | --- | --- | --- | --- | --- | --- | --- |
| 1 | 1550 | 1230 | 4760 | 3 | 0.30 | 284 | 23 |
| 2 | 1340 | 615 | 3160 | 3 | 0.30 | 437 | 11 |
| 3 | 2310 | 1540 | 4960 | 3 | 1.00 | 489 | 29 |
| 4 | 1210 | 588 | 3870 | 4 | 0.78 | 668 | 19 |
| 5 | 1710 | 1100 | 5530 | 3 | 0.20 | 500 | 7 |

CDVA = Corrected-distance-visual-acuity; CCT = Central corneal thickness; DM = Descemet membrane; MCT = Maximal corneal thickness at DM detachment site; PDL = Pre-Descemet’s layer/Dua’s layer
